# Prostate cancer risk prediction using polygenic hazard scores in Norwegian populations

**DOI:** 10.64898/2026.01.07.26343583

**Authors:** Andrew H. Morris, Bayram C. Akdeniz, Sigve Nakken, Alexey Shadrin, Anders M. Dale, Ole A. Andreassen, Eivind Hovig, Tyler M. Seibert, Oleksandr Frei

## Abstract

More accurate risk prediction is needed for prostate cancer (PCa) to identify individuals at greatest risk of early-onset and clinically significant disease. Current screening paradigms, such as prostate-specific antigen screening, pose risks due to over-diagnosis and over-treatment of indolent disease. Polygenic hazard scores (PHS) can predict age-at-diagnosis of PCa and are being tested in prospective clinical screening trials. We assessed the performance of the latest PHS model for PCa (PHS601) in two Norwegian population-based cohort studies (N = 14,688 and N = 2,850) for predicting age-at-diagnosis of PCa and aggressive PCa. In a subset with whole-genome sequencing (N = 503), we directly compared PHS601-based stratification with screening for rare pathogenic variants. PHS601 effectively stratified participants by risk in both cohorts for both PCa (HR_80/20_ = 5.74, 95% CI [5.06, 6.45] and 7.79 95% CI [5.70, 10.59]) and aggressive PCa (HR_80/20_ = 4.60, 95% CI [3.19, 6.45] and 3.14 95% CI [1.49, 6.63]). Among individuals with whole-genome sequencing, the top 1.8% of PHS values conferred 8.8-fold higher risk than the median (HR = 8.78 [5.00, 14.40]), exceeding the risk associated with HOXB13 pathogenic variants (HR = 3.77 [1.75, 8.11]; 1.8% carrier frequency). This study provides the first external validation of PHS601 in independent Norwegian population-based cohorts and, within a WGS subset, a direct comparison with rare variant screening, supporting context-specific use of genotyping-based risk stratification in population screening.

## 1 Introduction

Prostate cancer (PCa) is the second most commonly diagnosed cancer among men worldwide and a leading cause of cancer-related mortality^1^. Although early detection can reduce deaths from PCa, population screening with prostate-specific antigen (PSA) is limited by false positives, overdiagnosis, and overtreatment, particularly of indolent disease^2, 3^. Therefore, there is a clinical need to identify men at elevated risk of clinically significant or metastatic PCa to focus early screening and downstream diagnostics (e.g., MRI), while minimizing harms.

PCa is highly heritable (∼42%)^4^ with risk influenced by rare pathogenic variants (RPVs) as well as common variants. RPVs, such as in BRCA1, BRCA2, and HOXB13, confer large increases in risk, but occur at low frequencies in the general population^5^. Polygenic models combine many common variants with small effects to estimate inherited risk. Furthermore, polygenic hazard scores (PHS) use time-to-event modeling, which allows for prediction of age at diagnosis^6–8^. The latest age-of-onset polygenic hazard score, PHS601, has been integrated with family history and genetic ancestry to form P-CARE (Prostate CAncer integrated Risk Evaluation)^9^, which is being implemented in a clinical trial of precision prostate cancer screening in the United States Department of Veterans Affairs healthcare system (Clinicaltrials.gov NCT05926102, registered July 3rd, 2023).

PHS601 was developed in the US Million Veterans Program, a large, multi-ethnic cohort, and showed robust replication in the PRACTICAL consortium, but independent validation in population-based cohorts is lacking^9^. In addition, direct within-cohort comparisons of PHS-based stratification versus WGS-based rare pathogenic variant (RPV) screening are limited. Here, we performed external validation of PHS601 in two Norwegian cohorts: a pop-ulation study (Hordaland Health Studies; HUSK) and a clinical cohort enriched for family history of cancer (Norwegian Familial Cancer Study; NFCS). We evaluated whether PHS601 stratifies risk of PCa and aggressive PCa. We also leveraged whole-genome sequencing to compare PHS-based risk stratification with RPV carrier status. Our aim was to establish the performance of PHS601 for age-specific risk prediction in independent populations and to evaluate the relative utility of PHS versus RPV screening for genomically-informed cancer screening.

## 2 Methods

### 2.1 Study Population

This study evaluated two Norwegian cohorts that have not been included in prior PHS601 model development or validation (Table 1). The first cohort was the Hordaland Health Studies (Helseundersøkelsene i Hordaland; HUSK) with 35,515 participants. This cohort was recruited from Rogaland county in western Norway in two waves. The first wave was in 1992-1993 and the second wave was in 1997-1999. Analyses were restricted to the 14,688 male participants in HUSK. The second cohort was the Norwegian Familial Cancer Study (NFCS) with 15,769 participants^10^. Participants in this cohort were recruited in 1980-1995 from all of Norway via self-report based on over-representation of cancer in their families. Analyses were restricted to the 2,850 male participants in NFCS.

**Table 1:** Median and interquartile range (Q1, Q3) of key age and demographic variables for each cohort. Age at first follow-up is not available for the NFCS cohort. Age at aggressive diagnosis is restricted to aggressive cases. Age at last follow-up is restricted to controls (censored participants).

|  | HUSK | NFCS |
| --- | --- | --- |
| Birth Year | 1953 (1950, 1955) | 1941 (1934, 1950) |
| Recruitment Period | 1992–1999 | 1980–1995 |
| Age at First Follow-up | 58.8 (56.0, 63.2) | — |
| Age at Diagnosis | 66.2 (61.8, 70.1) | 64.0 (57.0, 71.0) |
| Age at Aggressive Diagnosis | 67.5 (62.9, 74.9) | 66.5 (57.2, 74.0) |

### 2.2 Characterization of PCa Cases

In both cohorts, PCa cases were defined as participants with ICD10 code C61. Diagnosis date was the first instance of this ICD10 code in the Cancer Registry of Norway, which has been found to be 98.8% complete^11^. Age was used as the time scale, with follow-up from birth to age at diagnosis (event) or age at last follow-up (censoring). Deaths prior to prostate cancer diagnosis were treated as censoring events. Aggressive disease was defined according to metastasis codes 1, 2, 4, 6, and 7 in the Norwegian Cancer Registry, corresponding to disease that extends to lymph nodes inside or outside the same body region, organ metastasis outside the body region, and microscopic or macroscopic ingrowth into neighboring structures. Progression, relapse, and metastases are required to be reported by the Norwegian Cancer Registry, supporting completeness of the aggressive phenotype ascertainment.

### 2.3 Genotyping and PHS calculation

#### 2.3.1 Genotyping

HUSK participants were genotyped at deCODE Genetics (https://www.decode.com/) in two batches (10,083 and 25,432 individuals in batch 1 and 2, respectively) using a customized Illumina GSA v3 array (687,316 genotyped variants). Prior to phasing and imputation, genotyping data were QCed separately for each batch using plink2^12^. QC steps for vari-ants included removal of indels, non-ATGC, strand-ambiguous, poorly-genotyped (plink2’s option: –geno 0.05), non-autosomal and rare (–maf 0.05) variants as well as variants with extreme deviation from Hardy-Weinberg equilibrium (–hwe 1E-50 midp keep-fewhet). Chromosomal strand and allele order for the remaining variants were aligned based on the HGDP + 1KG reference panel from gnomAD^13^, and variants with alleles that were not possible to align to the reference as well as variants with MAF deviating from the reference by more than 30% were removed. Resulting genotypes from two batches were merged. Variants with high missingness (–geno 0.02) and individuals with poor genotyping rate (–mind 0.05) were removed from the merged data. The combined dataset was then phased and imputed with Beagle 5.4 (version 27May24.118) using the HGDP + 1KG reference panel. The resulting dataset contained 35,476 participants and 76,413,521 autosomal variants.

For the NFCS cohort, genomic DNA was extracted from peripheral blood samples using the DNeasy Blood & Tissue Kit (Qiagen, Germantown, MD, USA), following the manufacturer’s protocol, and analyzed using the Illumina OmniExpress 24 v1.1 chip at deCODE genetics (Reykjavik, Iceland). The genotyped dataset consisted of 713,014 SNPs. Genotype data were imputed using the HRC v1.1 reference panel following the same imputation protocol as The Norwegian Mother, Father and Child Cohort^14^. After imputation, the dataset consisted of 39,111,193 autosomal variants.

#### 2.3.2 Harmonization of alleles and calculation of PHS

The list of variants used by the PHS601 model, and their effect sizes, were acquired by contacting the authors^9^. Chromosome X imputation was not available in our genotype data so the 21 SNPs in the original model on chromosome X were removed leaving 580 SNPs. Imputed genotype data in each cohort and model variants were matched by genomic position and alleles, resulting in 519 variants (in HUSK) and 557 variants (in NFCS) used for PHS computation. Strand-ambiguous variants were retained, after verifying that no strand flips were present among non-strand ambiguous SNPs in either of the two cohorts. Polygenic hazard scores were calculated for each cohort using plink--score^12^. Missing genotypes contributed an amount proportional to the imputed allele frequency.

### 2.4 Whole-genome sequencing

#### 2.4.1 Sequencing

The whole-genome sequencing data were generated at deCODE Genetics on samples from 2,576 participants from the NFCS cohort as described previously^15^. The samples were sequenced using Illumina HiSeqX and NovaSeq sequencing platforms with a total of 8,034 lanes, and all samples were prepared without PCR. Duplicated samples were discarded based on sequencing yield and only samples with a genome-wide average coverage of 20× and higher were included. read_haps was used to detect and remove contaminated samples^16^. The average genome-wide sequencing coverage was 37.2X (SD = 6.3; minimum = 20.1X; maximum = 98.5X) and joint variant calling was performed using GraphTyper (v.2.7.5)^17^.

#### 2.4.2 Variant calling and classification of RPVs

Identification of RPVs in the WGS dataset was performed using the Cancer Predisposition Sequencing Reporter – CPSR v2.2.0^18^. Specifically, we used CPSR to screen WGS variant calls for heterozygous and homozygous SNVs/InDels in high-penetrance prostate cancer genes, including BRCA1, BRCA2, ATM, PALB2, CHEK2, HOXB13, MLH1, MSH2, MSH6, PMS2, TP53, and EPCAM. Coding variants in these genes classified as pathogenic according to ClinVar (release 2025_03) were considered RPVs (Table S1). Of note, the HOXB13 G84E variant (rs138213197) was included in PHS601 and was also identified among the RPVs in whole-genome sequenced participants. All participants carrying the G84E mutation in WGS data also carried rs138213197 in the imputed genotype data.

### 2.5 Statistical Analysis

#### 2.5.1 Polygenic hazard score performance evaluation

Polygenic hazard scores were evaluated in both cohorts using Cox proportional hazards models using the coxph function from the survival package in R^19^. Polygenic hazard scores were centered and scaled prior to model fit. Performance of each model is reported as HR_SD_, which is the increase in risk for a one-SD increase in PHS, HR_80/20_, which is the hazard ratio between the top 20% and the bottom 20% of PHS, HR_80/50_, which is the hazard ratio between the top 20% and the median PHS (30th to 70th percentile), HR_95/50_, which is the hazard ratio between the top 5% and the median PHS (30th to 70th percentile), and Harrell’s concordance index (C-index). For these metrics, 95% confidence intervals were generated using 999 bootstrap iterations. All Cox models were evaluated for the proportional hazards assumption using cox.zph and by inspecting plots of the Schoenfeld residuals versus time^20^. For models that did not meet these assumptions, we performed sensitivity analysis with time-dependent covariates and a linear time transform (i.e., PHS ⋅ age) for the time-dependent PHS with the tt function.

#### 2.5.2 Rare pathogenic variant screening and comparison to polygenic risk

We assessed the effect of RPVs on PCa risk using WGS data from the whole-genome sequenced subset of the NFCS cohort. We first tested whether RPVs in any of the twelve genes were enriched among cases versus controls using a one-sided Fisher’s exact test (one for each gene plus one for any-gene carrier status for a total of thirteen tests). Genes with significant enrichment (Benjamini-Hochberg adjusted p < 0.05) were then modeled in Cox proportional hazards analyses to estimate HR between carriers and non-carriers and C-index. To compare the RPV risk stratum to PHS, we restricted analyses to participants with both WGS and genotyping data, refitting the PHS Cox model in this subset and calculating HR for a PHS percentile stratum matching the RPV carrier frequency.

#### 2.5.3 Comparison with Norwegian Cancer Registry data

To evaluate how generalizable our results were to the population of Norway, we compared the cumulative incidence in each cohort for both PCa (any stage) and for aggressive PCa to the cumulative incidence from the Cancer Registry of Norway (using the same ICD10 code and metastasis codes as before). We used annualized age-specific incidence rate binned by 5-year age groups for the years 2014-2017, as previously reported^21^. To convert the 5-year incidence rate into cumulative incidence, we fit a log-quadratic model (for PCa) and a log-linear model (for aggressive PCa) to the average incidence for each 5-year age group. We then predicted the annualized incidence from this model and calculated the cumulative incidence from the cumulative sum of cases. We compared these to the cumulative incidence derived from the Cox proportional hazards model for each phenotype (PCa or aggressive PCa) in each cohort. We then stratified the cumulative incidence curves by HRs from the PHS Cox proportional hazards model fit to the HUSK data.

## 3 Results

### 3.1 PHS601 stratified PCa risk in two Norwegian cohorts

Participants with higher genetic risk scores in PHS601 showed greater risk of early diagnosis in both Norwegian cohorts. In the HUSK cohort (1650 cases and 13,038 controls), each SD increase in PHS was associated with an 87% greater risk of PCa (HR_SD_ = 1.87, 95% CI [1.79, 1.95]), with individuals in the top quintile having over 5 times the risk of those in the bottom quintile (HR_80/20_ = 5.74, 95% CI [5.06, 6.45]; Table 2 and Figure 1). Results were similar in the NFCS cohort (339 cases and 2511 controls), where each SD increase in PHS conferred a twofold greater risk (HR_SD_ = 2.09, 95% CI [1.87, 2.33]; Table 2), with HR_80/20_ = 7.79 (95% CI [5.70, 10.59]; Table 2 and Figure 1). Cumulative incidence in the HUSK cohort was consistent with national Cancer Registry data, though the NFCS cohort showed elevated baseline incidence relative to national data (Figure S1).

**Figure 1:**
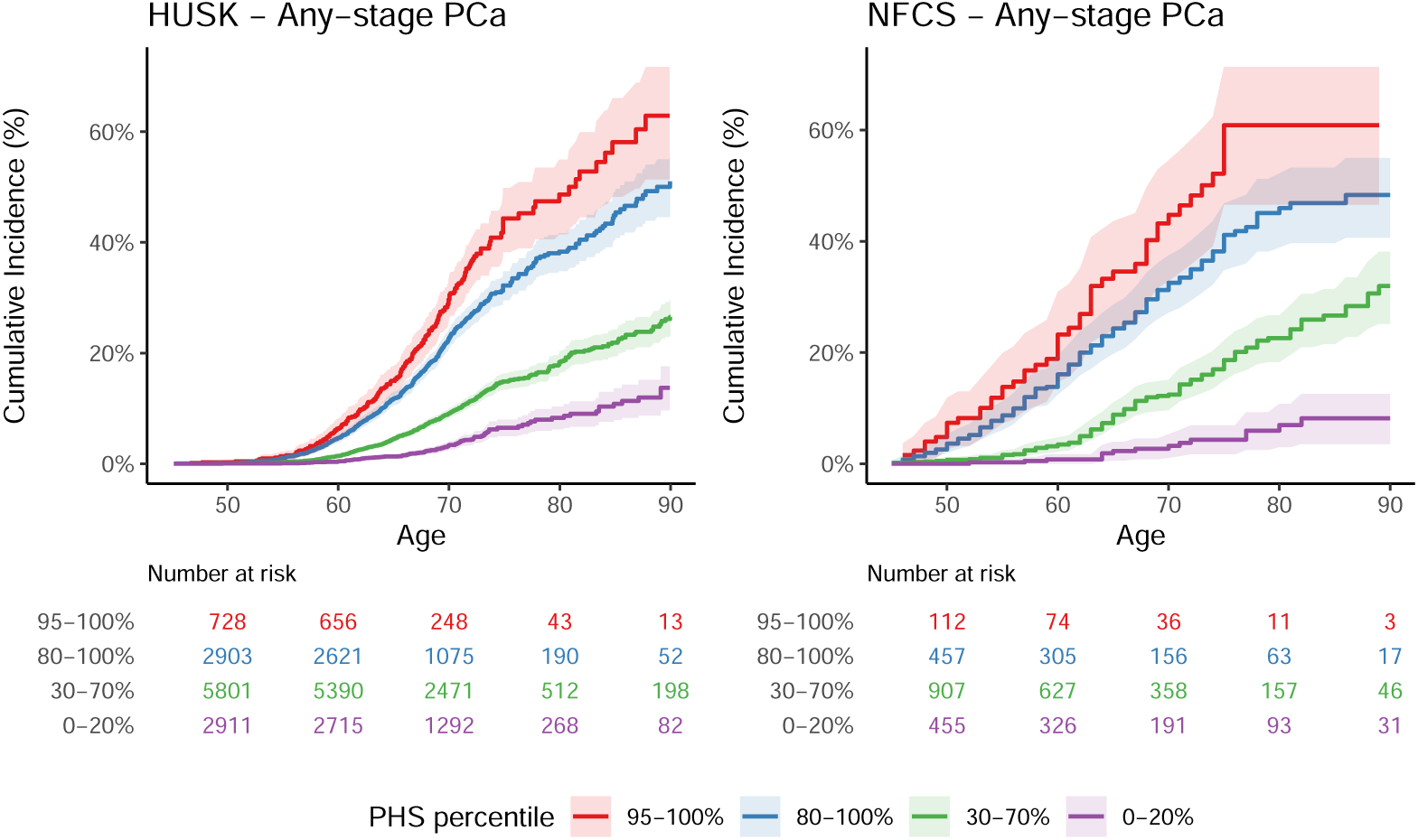
Cumulative incidence of PCa by age and stratified by centile ranges of PHS for the HUSK and NFCS cohorts. Time of event is age at diagnosis of PCa (any stage). Controls were censored at age of last observation. Error ribbons are the 95% confidence interval.

**Table 2:** HRs and C-Index for the any-PCa and aggressive-PCa models. HR_SD_ is the increase in HR for a one-standard-deviation increase in PHS. HR_80/20_ is the HR between the top quintile and the bottom quintile of PHS. HR_80/50_ is the HR between the top quintile and the median PHS (30-70th percentile). HR_95/50_ is the HR between the top 5% and the median PHS (30-70th percentile). 95% confidence intervals are based on 999 bootstraps.

| Cohort | Cases | $HR_{SD}$ | $HR_{80/20}$ | $HR_{80/50}$ | $HR_{95/50}$ | C-Index |
| --- | --- | --- | --- | --- | --- | --- |
| **Any-stage PCa** |  |  |  |  |  |  |
| HUSK | 1650 | 1.87 (1.79, 1.95) | 5.74 (5.06, 6.45) | 2.49 (2.33, 2.65) | 3.96 (3.59, 4.36) | 0.69 (0.68, 0.70) |
| NFCS | 339 | 2.09 (1.87, 2.33) | 7.79 (5.70, 10.59) | 2.90 (2.45, 3.42) | 5.01 (3.84, 6.48) | 0.72 (0.69, 0.75) |
| **Aggressive PCa** |  |  |  |  |  |  |
| HUSK | 200 | 1.73 (1.52, 1.95) | 4.60 (3.19, 6.45) | 2.22 (1.83, 2.64) | 3.32 (2.50, 4.33) | 0.65 (0.61, 0.70) |
| NFCS | 46 | 1.51 (1.15, 1.97) | 3.14 (1.49, 6.63) | 1.81 (1.23, 2.68) | 2.45 (1.37, 4.49) | 0.64 (0.55, 0.73) |

PHS effects were strongest at younger ages in both cohorts. Schoenfeld residuals tests revealed a negative trend in HR with age (HUSK: χ^2^ = 10.21, p = 0.001; NFCS: χ^2^ = 9.98, p = 0.002; Figure S2) confirmed with a time-varying Cox model with a linear age interaction, which showed that genetic risk declined by 1.1-1.6% per year (Table S2). Given the weak magnitude of this interaction and high residual variability, we present proportional hazards models as the primary analysis; the age attenuation finding is discussed below.

### 3.2 PHS601 was associated with aggressive PCa in Norwegian cohorts

In addition to evaluating the performance of PHS601 in stratifying individuals based on risk of PCa at any stage, we were also interested in the association of PHS601 with aggressive disease. In the HUSK cohort (200 aggressive cases and 14,488 controls), each SD increase in PHS was associated with a 73% greater risk of aggressive disease (HR_SD_ = 1.73, 95% CI [1.52, 1.95]), while individuals in the top quintile of PHS had 4-fold the risk of those in the bottom quintile (HR_80/20_ = 4.60, 95% CI [3.19, 6.45]; Table 2 and Figure 2). Similarly, in the NFCS cohort (46 aggressive cases and 2,804 controls), each SD increase in PHS conferred a 51% greater risk of aggressive disease (HR_SD_ = 1.51, 95% CI [1.15, 1.97]), with HR_80/20_ = 3.14 (95% CI [1.49, 6.63]; Table 2 and Figure 2). Proportional hazards assumptions were not violated in either cohort based on statistical tests and weighted residual plots (HUSK: χ^2^ = 0.23, p = 0.63; NFCS: χ^2^ = 1.73, p = 0.19; Figure S2).

**Figure 2:**
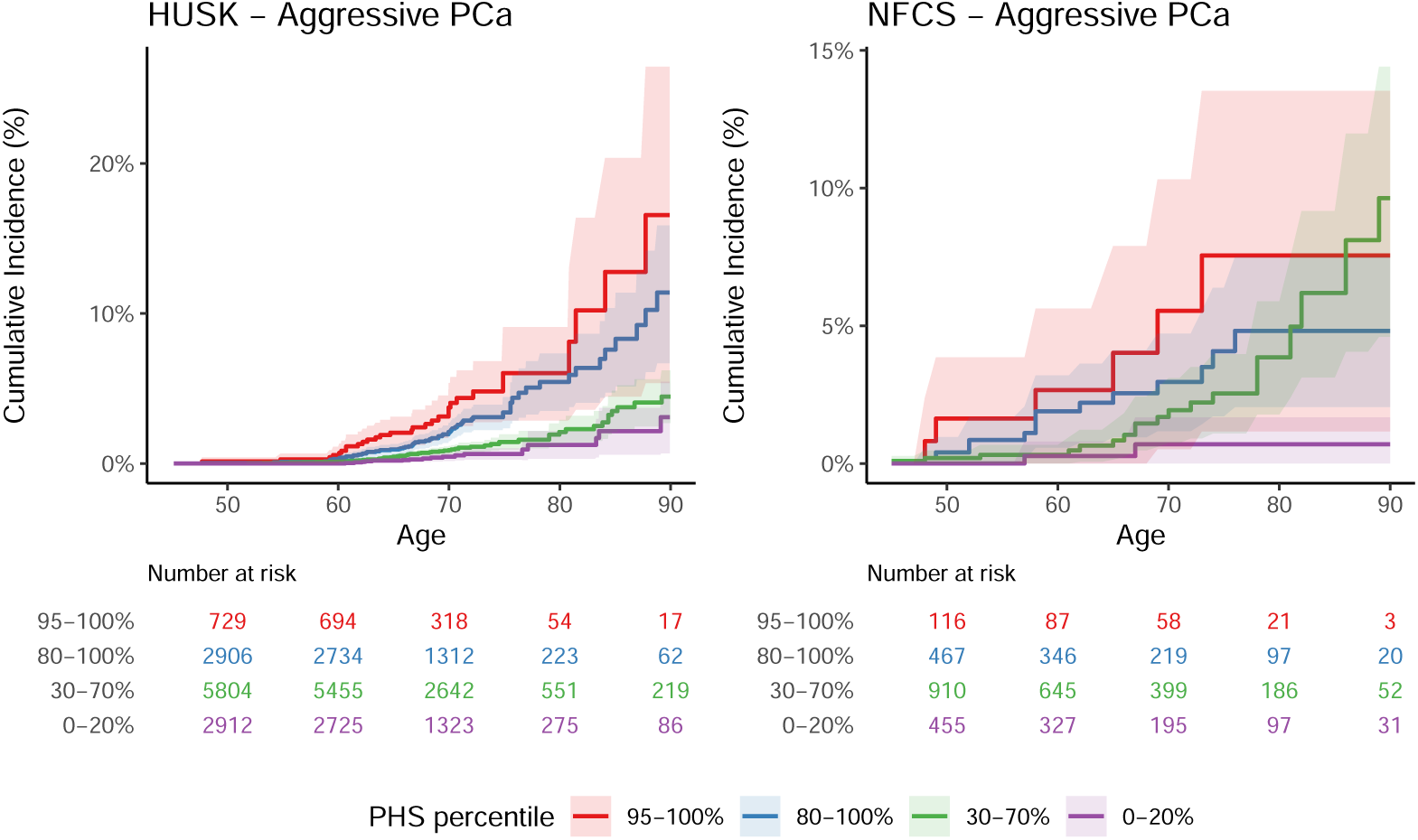
Cumulative incidence of aggressive PCa by age and stratified by centile ranges of PHS for the HUSK and NFCS cohorts. Time of event is age at diagnosis of aggressive PCa. Controls were censored at age of last observation. Error ribbons are the 95% confidence interval.

### 3.3 PHS provided stronger population-level stratification than RPV screening across known PCa predisposition genes

We compared RPVs and PHS in predicting PCa risk in the NFCS cohort. Analyses were restricted to 503 participants with both WGS and genotyping (138 cases and 365 controls) of whom 27 carried a predisposing mutation in one of the twelve target genes. Of the twelve potentially predisposing genes, only HOXB13 had an enrichment in the number of pathogenic variants among PCa cases compared to controls (OR = 9.65, 95% LCB = 2.23, adj. p = 0.03; Table S3); subsequent survival models therefore focus on HOXB13 as the only gene with evidence of enrichment.

In the WGS subset, a one-SD increase in PHS predicted a 92% higher risk of PCa (HR = 1.92, 95% CI [1.66, 2.22], p < 0.001; Figure 3), while HOXB13 carriers had a 3.8-fold greater risk than non-carriers (HR = 3.77, 95% CI [1.75, 8.11], p < 0.001). Similarly, the PHS model had a C-index of 0.702 (SE = 0.022) whereas the HOXB13 model had a C-index of 0.519 (SE = 0.009). HOXB13 pathogenic variants occurred in 1.8% of participants. To enable direct comparison with this risk stratum, we evaluated individuals in the top 1.8% of the PHS distribution against the median. This high-risk PHS stratum demonstrated 8.8-fold higher risk (HR_98.2/50_ = 8.78, 95% CI [5.00, 14.40]), exceeding the risk conferred by HOXB13 pathogenic variants (HR = 3.77).

**Figure 3:**
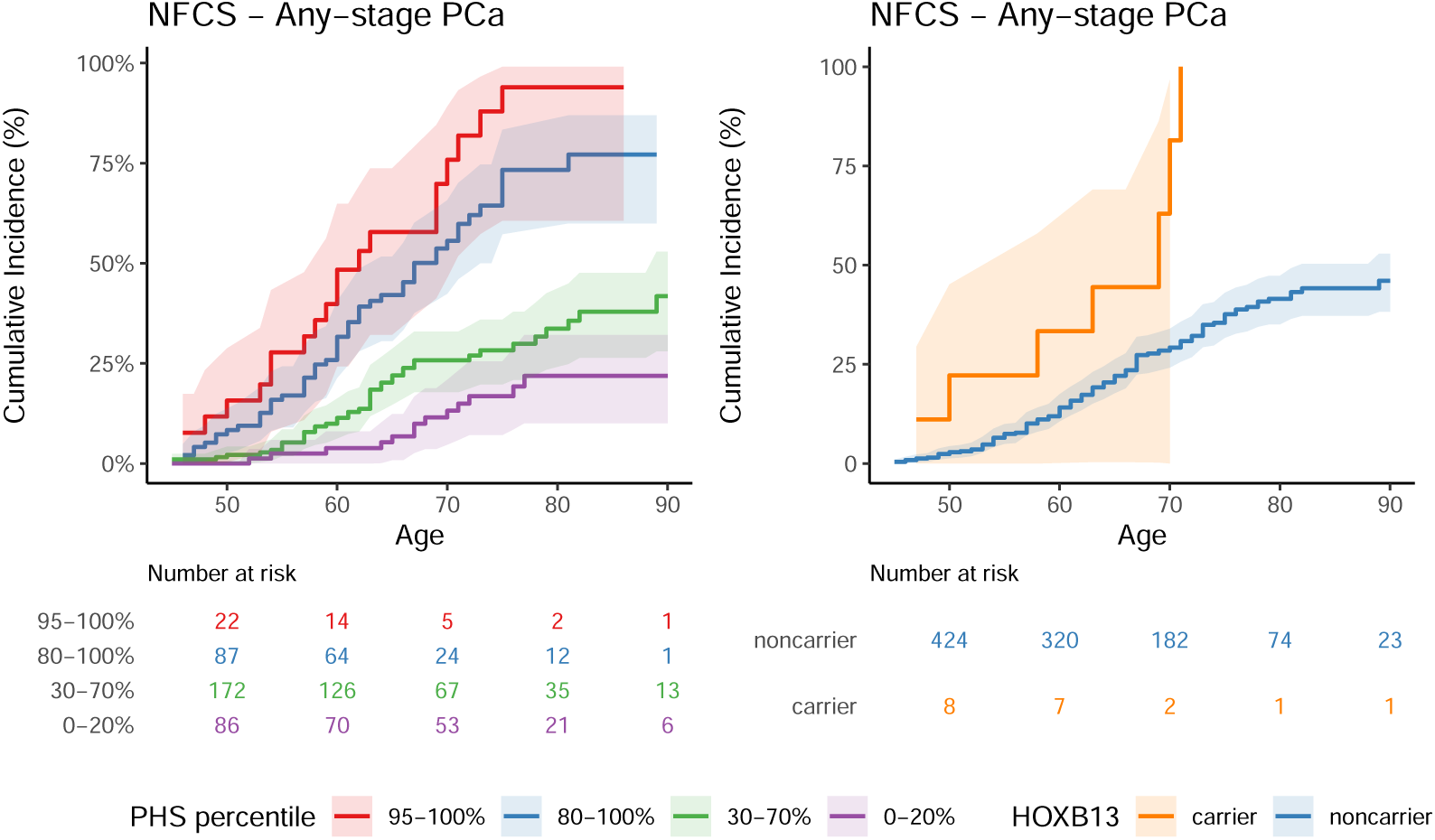
Cumulative incidence of PCa by age for the whole-genome sequenced subsample of the NFCS cohort stratified by either centile ranges of PHS or HOXB13 carrier status. Time of event is age at diagnosis of PCa (any stage). Controls were censored at age of last observation. Error ribbons are the 95% confidence interval.

## 4 Discussion

We provide the first external validation of PHS601 in independent European population-based cohorts, demonstrating that the time-to-event genetic risk stratification of PHS601 generalizes across healthcare systems and populations with different baseline risk profiles, screening practices, and genetic backgrounds. The magnitude of risk stratification in HUSK (HR_80/20_ = 5.74, 95% CI [5.06, 6.45]) was consistent with PHS601 replication in the PRACTICAL consortium (COSM: HR_80/20_ = 9.18, 95% CI [6.66–12.93]; ProtecT: HR_80/20_ = 5.67, 95% CI [4.75, 6.73])^9^, supporting robust cross-population transferability of the model to independent European cohorts outside the development and prior validation populations. To illustrate the clinical magnitude of this risk stratification: by age 75, men in the top PHS quintile had an estimated cumulative PCa incidence of approximately 30%, compared with ∼8% for those in the bottom quintile (Figure 4), a ∼4-fold absolute risk difference.

**Figure 4:**
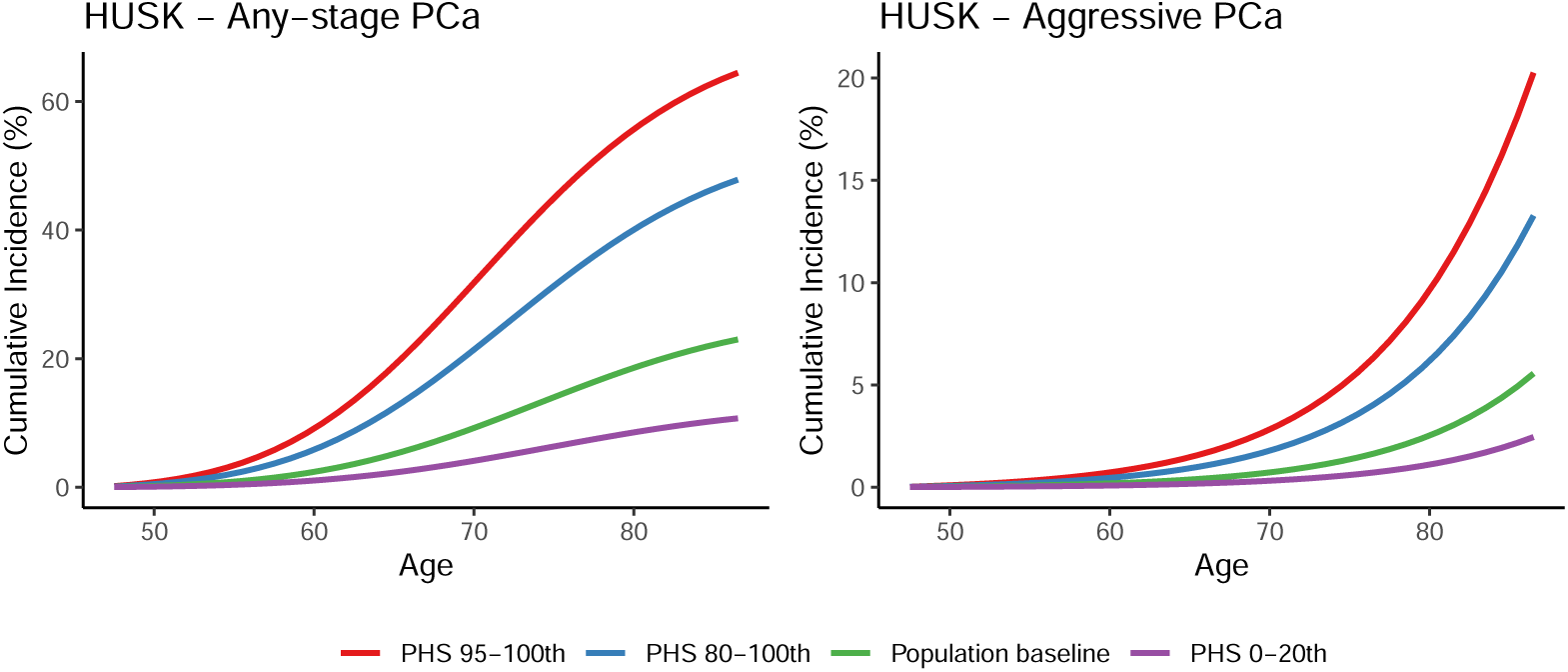
Predicted cumulative incidence of PCa and aggressive PCa in the general population of Norway based on data from the Norwegian Cancer Registry. The population baseline reflects PCa cases in Norway from 2014-2017^21^. Incidence curves are stratified by the HR of PHS centiles from the PHS601 model fit to the HUSK data. The decline in incidence after age 70 observed for any-stage PCa is likely due to decreased screening rather than a true reduction in incidence.

PHS601 stratified absolute risk of aggressive PCa in both cohorts, despite being trained on any-stage disease, confirming prior associations of PHS601 with metastatic and clinically-significant disease^9, 22^, though risk stratification for aggressive PCa was weaker than for any-stage PCa (HUSK: HR_80/20_ for aggressive = 4.60, 95% CI [3.19, 6.45] versus any-stage PCa = 5.74, 95% CI [5.06, 6.45]). This study assessed risk from birth in the general population, which is the appropriate framing for a pre-screening context, since the clinically actionable question is who among undiagnosed men carries elevated inherited risk of aggressive disease. A case-only analysis would inappropriately condition on prior diagnosis^23^. Once a diagnosis is established, germline risk is superseded by superior instruments for predicting progression, including PSA trajectory, Gleason grade, MRI staging, and somatic genomic classifiers such as Decipher and Oncotype DX. Critically, PHS serves the screening decision rather than the treatment decision. High-PHS men benefit from earlier and more intensive surveillance, while low-PHS men are candidates for reduced or deferred screening — directly addressing the overdiagnosis harms that limit current PSA-based screening paradigms. The clinical relevance of PHS for aggressive disease is further supported by evidence that earlier PCa diagnosis independently confers elevated lifetime risk of metastatic progression regardless of initial grade^24^. High-PHS men are enriched for early-onset disease and therefore carry elevated cumulative metastatic risk.

In addition to testing the performance of PHS, we screened twelve known PCa predisposition genes and found that, in this dataset, genotyping-based PHS provided stronger population-level risk stratification than WGS-based RPV screening. Only HOXB13 showed enrichment among cases, and even this variant performed substantially worse than PHS. A much larger percentage of the population is affected by a high risk score than is affected by currently known RPVs—high polygenic risk affects 50x more people than BRCA2 mutations^5^—which means genetic risk scoring may be more useful at the population level. In this context, HOXB13 is best interpreted as a risk-associated variant rather than a deterministic disease marker, and its low carrier frequency limits utility for broad population-level identification of at-risk men. Notably, genotyping with imputation successfully identified all HOXB13 carriers subsequently confirmed by WGS, suggesting that for population-scale PCa screening, a single genotyping array can simultaneously generate PHS and capture the only known high-effect PCa variant evaluated here.

Even though HOXB13 did not discriminate as well as the PHS model, clinical interventions may differ between individuals whose phenotype arises from rare deleterious mutations versus those whose phenotype arises from the interaction of environment with the normal array of segregating genes of minor effect. For example, individuals with BRCA mutations may benefit from PARP inhibitors^25^. Furthermore, RPVs and PHSes have different implications for genetic counseling, particularly regarding the risk to offspring in the case of RPVs. Therefore, there remains value in screening for these rare variants in appropriate clinical contexts.

We observed a weak but significant negative trend in the HR of PHS with age in two independent cohorts such that the strongest effect of genetic risk was at the youngest ages (Figure S2). In time-varying models, this corresponded to an estimated decline in genetic effect size of approximately 1.1-1.6% per year (Table S2). This age-dependent attenuation of genetic risk is consistent with observations from other complex diseases and may reflect several biological processes. First, as individuals age, the cumulative burden of environmental exposures, somatic mutations, and stochastic cellular processes may increasingly dominate over germline genetic variation in determining disease risk^26, 27^. Second, competing risks of death from other causes may differentially affect high-risk individuals, leading to a selected survivor population at older ages. Stronger genetic effects at younger ages have been well-documented for BRCA1/2 mutations in breast cancer^28^ and across multiple complex diseases^26^. These findings identify the age range where genetic risk stratification may be most actionable for prevention and early detection.

This study has several limitations. We did not have access to family history data or prostate-specific antigen measurements for all participants. While family history provides additional information beyond PHS601 at the population level, and is included as an independent predictor in the integrated P-CARE model^9^, future work should explore integration of PHS with other clinical risk factors and with social determinants of health, lifestyle factors, and environmental exposures^29–31^. The NFCS cohort was enriched for family history of cancer and is not representative of the general Norwegian population (Figure S1); absolute risk estimates were therefore derived from the population-based HUSK cohort. Whole-genome sequencing was available only in a subset of NFCS, which reduced power for very rare variant analyses and does not preclude additional rarer variants with larger effects; therefore, the PHS-versus-RPV comparison should be interpreted as a comparison of observed effect sizes in this dataset. Future implementation studies should also evaluate performance and access equity across diverse ancestry and healthcare groups to avoid widening disparities. The strong risk stratification observed here provides a foundation for prospective trials of risk-stratified screening in Norway, but cost-effectiveness analyses will be needed before clinical implementation to identify optimal screening thresholds and age ranges.

## 5 Conclusions

We demonstrated that PHS601 performs well in independent, population-based cohorts, providing strong risk stratification for both prostate cancer and aggressive disease. This represents the first external validation in Norwegian population-based cohorts with a distinct healthcare system, baseline risk profile, and genetic background from the development and prior validation populations, and includes a direct within-cohort comparison of PHS and WGS-based rare variant screening. In this setting, genotyping-derived PHS601 provided stronger population-level risk stratification than screening across twelve known predisposition genes in WGS. Given that high polygenic risk affects far more individuals than known RPVs, a PHS-based screening paradigm has the potential for substantial population health impact. Lastly, the time-varying effect suggests PHS may have the greatest clinical utility at younger ages. These findings support the potential utility of PHS in a precision screening paradigm for PCa.

## Data Availability

The data that support the findings of this study are not publicly available due to patient privacy and ethical restrictions. Data were accessed through the Tjenester for Sensitive Data facilities owned by the University of Oslo under ethics approval by the Regional Committee for Medical Research Ethics South East Norway. Inquiries about data access should be directed to the Regional Committees for Medical and Health Research Ethics of Norway.

## Conflit of interest statement

Dr. Anders M. Dale is Founding Director, holds equity in CorTechs Labs, Inc. (DBA Cortechs.ai), and serves on its Board of Directors. Dr. Dale is the President of J. Craig Venter Institute (JCVI) and is a member of the Board of Trustees of JCVI. He is an unpaid consultant for Oslo University Hospital.

Dr. Ole Andreassen has received speaker fees from Lundbeck, Janssen, Otsuka, Lilly, and Sunovion and is a consultant to Cortechs.ai. and Precision Health.

Dr. Oleksandr Frei is a consultant to Precision Health.

## Acknowledgements

This work was supported by a grant from the Research Council of Norway (award #324499). The computations were performed on resources provided by Sigma2—the National Infrastructure for High-Performance Computing and Data Storage in Norway. This work was performed on the Tjenester for Sensitive Data (TSD) facilities, owned by the University of Oslo, operated and developed by the TSD service group at the University of Oslo IT-Department (UiO IT).

## Ethics statement

The study protocol was approved by the Norwegian Regional Ethics Committee (REK) Sør-Øst, IDs 2015/2382 (NFCS cohort) and 270326 (HUSK cohort).

---

**Figure S1:**
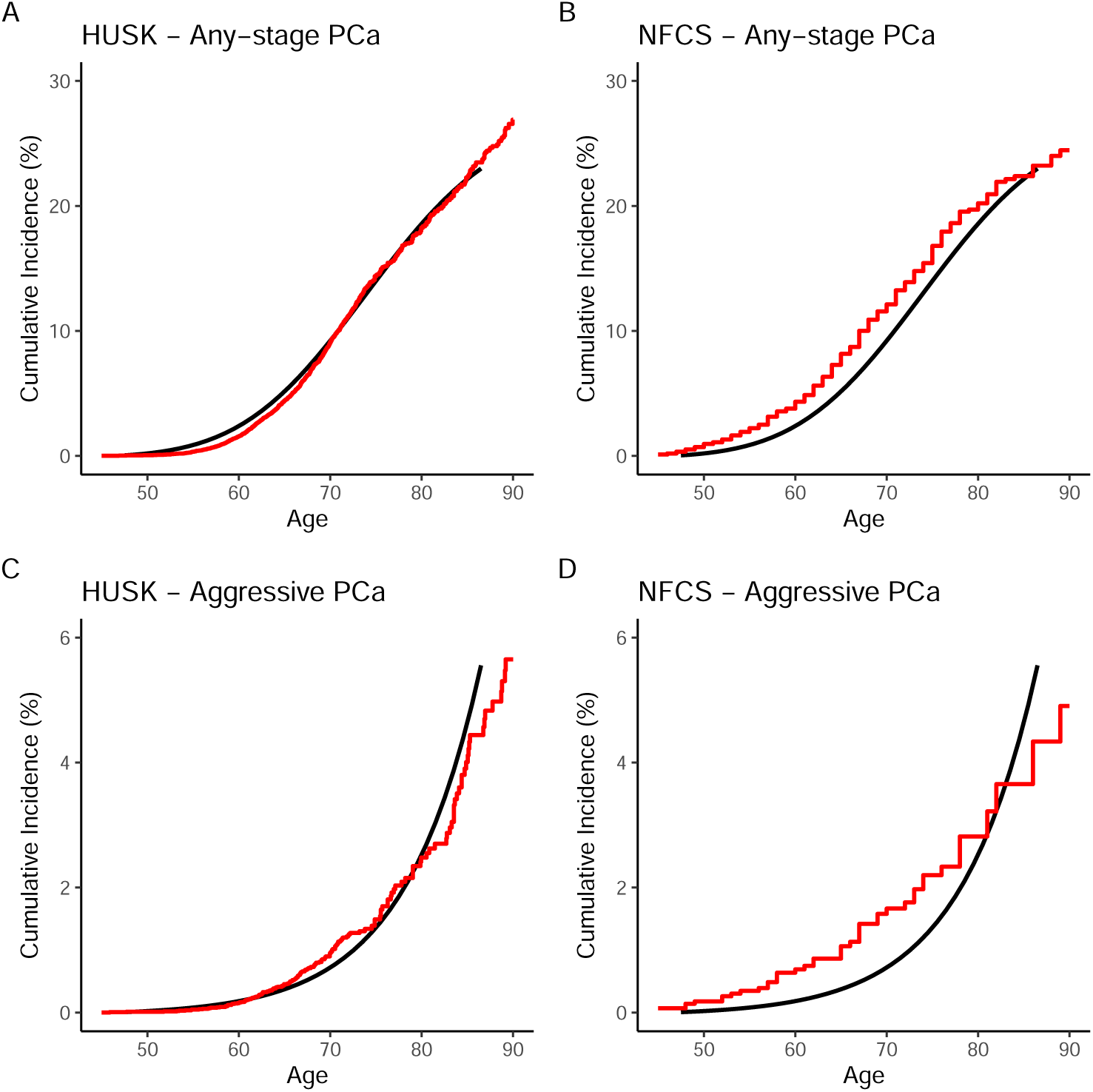
Comparison of cumulative incidence rates for PCa in two cohorts against national data from the Norwegian Cancer Registry. The red line represents the cumulative incidence observed in each cohort, while the black line depicts the overall cumulative incidence of PCa cases in Norway from 2014 to 2017^21^. Panels **(A)** and **(B)** show incidence rates for any-stage PCa in the HUSK and NFCS cohorts, respectively; panels **(C)** and **(D)** show incidence rates for aggressive PCa in the same cohorts.

**Figure S2:**
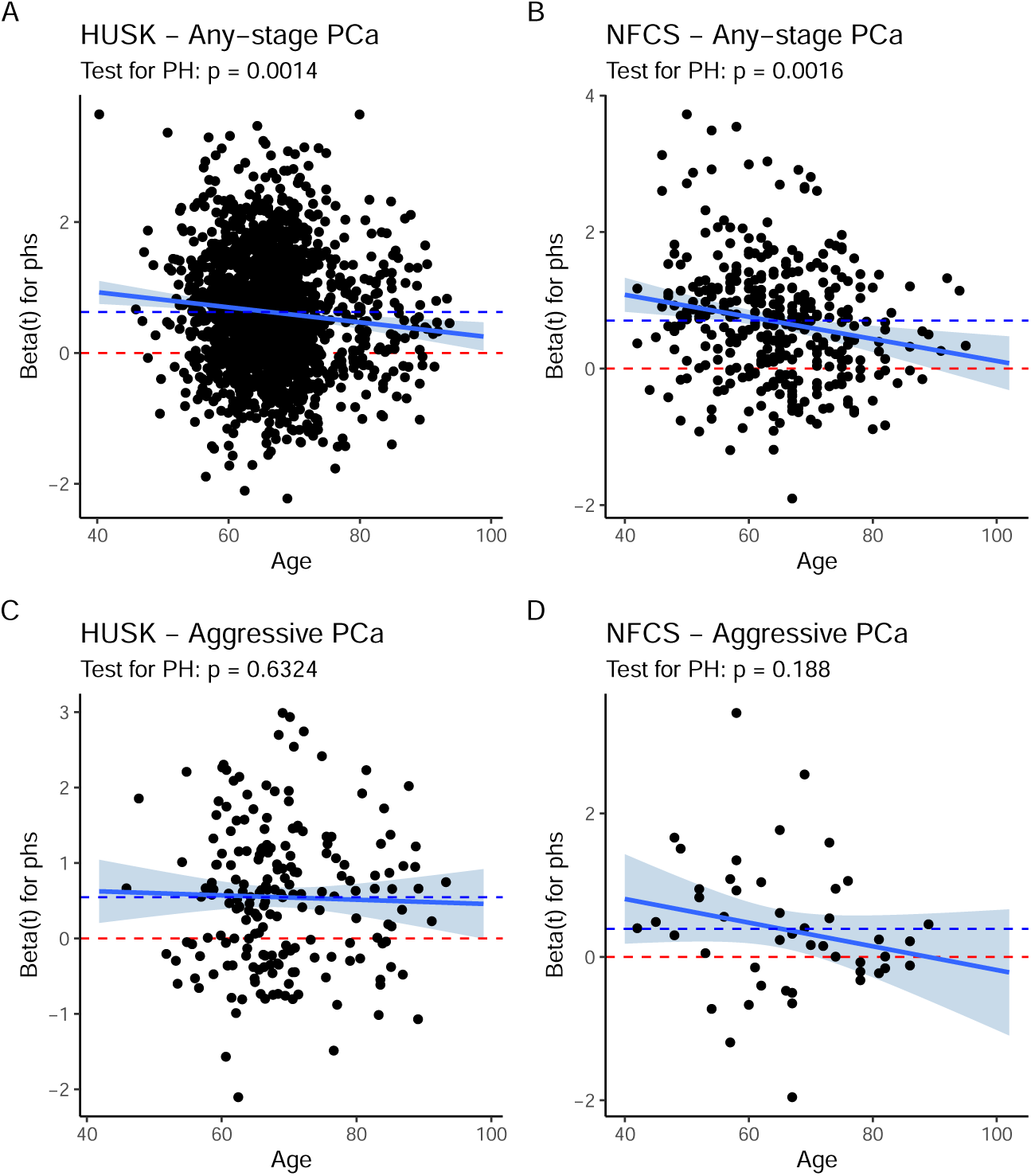
Assessment of the proportional hazards assumption for PHS using Schoenfeld residuals plotted against age. The solid blue line represents the estimated time-varying log hazard ratio from the extended Cox model with a linear time transform for PHS (95% confidence bands shown). The dashed blue line shows the log hazard ratio for PHS from the standard Cox proportional hazards model, and the dashed red line indicates a hazard ratio of 1.0 (log HR = 0). Divergence between the solid and dashed blue lines indicates violation of the proportional hazards assumption and time-dependency of the PHS effect. Panels **(A)** and **(B)** show results for any-stage PCa in the HUSK and NFCS cohorts, respectively; panels **(C)** and **(D)** show results for aggressive PCa in the same cohorts.

**Table S1:** All variants from genes that we investigated were classified as “Pathogenic” in ClinVar. N is the number of participants carrying this mutation.

| Symbol | Genomic Change | Alteration | dbSNP RSID | ClinVar Classification | N |
| --- | --- | --- | --- | --- | --- |
| ATM | 11:g.108272812C>CTG | p.His1082LeufsTer28 | rs761486324 | Pathogenic | 3 |
| ATM | 11:g.108272814TC>T | p.His1083ThrfsTer26 | rs767099464 | Pathogenic | 3 |
| BRCA1 | 17:g.43063930C>T | p.Arg1699Gln | rs41293459 | Pathogenic | 2 |
| BRCA1 | 17:g.43093974CT>C | p.Lys519ArgfsTer13 | rs80357662 | Pathogenic | 1 |
| BRCA1 | 17:g.43124096T>C | p.Met1? | rs80357287 | Pathogenic | 1 |
| BRCA2 | 13:g.32362595G>C | p.Trp2626Cys | rs80359013 | Pathogenic | 3 |
| BRCA2 | 13:g.32379440C>T | p.Gln2960Ter | rs80359140 | Pathogenic | 1 |
| CHEK2 | 22:g.28695868AG>A | p.Thr367MetfsTer15 | rs555607708 | Pathogenic | 3 |
| HOXB13 | 17:g.48728343C>T | p.Gly84Glu | rs138213197 | Pathogenic | 9 |
| MLH1 | 3:g.37004444C>T | p.Thr117Met | rs63750781 | Pathogenic | 1 |
| MLH1 | 3:g.37048973C>T | p.Arg687Trp | rs63751275 | Pathogenic | 1 |
| PALB2 | 16:g.23634953CA>C | p.Leu531CysfsTer30 | rs180177102 | Pathogenic | 1 |
| TP53 | 17:g.7673802C>T | p.Arg273His | rs28934576 | Pathogenic | 1 |
| TP53 | 17:g.7674217C>G | p.Arg249Thr | rs587782329 | Pathogenic | 1 |

**Table S2:**
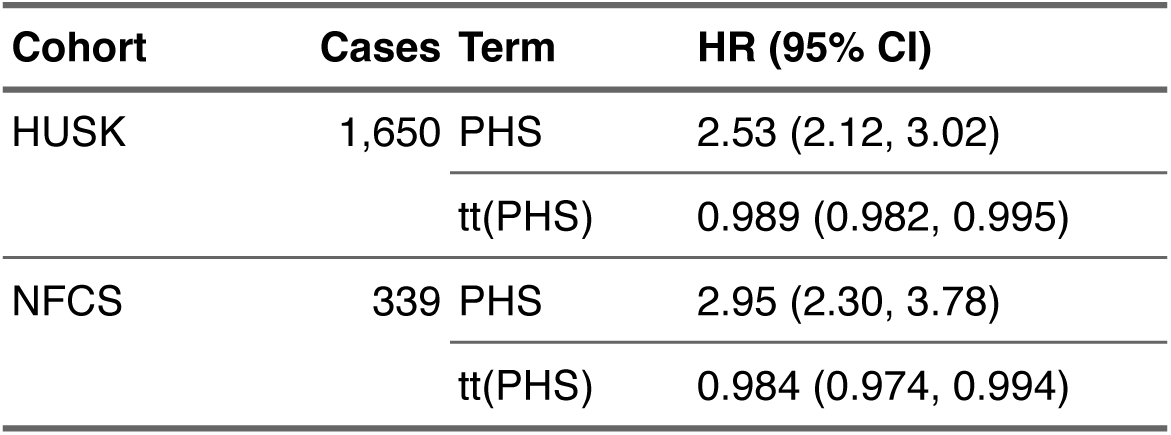
Hazard ratios and 95% confidence intervals from the extended Cox model for PHS with time-varying effects. The baseline PHS term represents the hazard ratio at age 40; the tt(PHS) term represents the multiplicative change in hazard ratio per additional year of age (linear time transform). Outcome is age at diagnosis of any-stage PCa.

**Table S3:** One-sided Fisher’s exact tests of whether the odds of carrying an RPV are greater among cases than controls in the NFCS cohort. P values adjusted using the Benjamini & Hochberg method. OR = Odds ratio, LCB = lower confidence bound.

| **RPV** | **OR (LCB)** | **P-value** | **Adj. P-value** |
| --- | --- | --- | --- |
| BRCA1 | 0.88 (0.03) | 0.720 | 1.000 |
| BRCA2 | 0.00 (0.00) | 1.000 | 1.000 |
| ATM | 0.00 (0.00) | 1.000 | 1.000 |
| PALB2 | 0.00 (0.00) | 1.000 | 1.000 |
| CHEK2 | 5.33 (0.41) | 0.180 | 0.800 |
| HOXB13 | 9.65 (2.23) | 0.002 | 0.029 |
| MLH1 | 0.00 (0.00) | 1.000 | 1.000 |
| MSH2 | 0.00 (0.00) | 1.000 | 1.000 |
| MSH6 | 0.00 (0.00) | 1.000 | 1.000 |
| PMS2 | 0.00 (0.00) | 1.000 | 1.000 |
| TP53 | 2.65 (0.07) | 0.470 | 1.000 |
| EPCAM | 0.00 (0.00) | 1.000 | 1.000 |
| Any | 1.89 (0.88) | 0.088 | 0.570 |

